# Effective vaccination strategies to control COVID-19 outbreak: A modeling study

**DOI:** 10.1101/2023.06.18.23291573

**Authors:** Youngsuk Ko, Kyong Ran Peck, Yae-Jean Kim, Dong-Hyun Kim, Eunok Jung

**Author notes:** **Address for Correspondence:** Eunok Jung, PhD Department of Mathematics, Konkuk University, Seoul, Korea.

## Abstract

**OBJECTIVES:** Three years following the start of the COVID-19 pandemic, the World Health Organization (WHO) declared COVID-19 a global health emergency of international concern. As immunity levels in the population acquired through past infections and vaccinations have been decreasing, booster vaccinations have become necessary to control new outbreaks. This study aimed to determine the most suitable vaccination strategy to control the COVID-19 surge.

**METHODS:** A mathematical model was developed to simultaneously consider the immunity levels induced by vaccines and infections, and employed to analyze the possibility of future resurgence and control using vaccines and antivirals.

**RESULTS:** As of May 11, 2023, a peak in resurgence is predicted to occur around mid-October of the same year if the current epidemic trend continues without additional vaccinations. In the best scenario, the peak number of severely hospitalized patients can be reduced by 43% (480) compared to the scenario without vaccine intervention (849). Depending on the outbreak trends and vaccination strategies, the best timing for vaccination in terms of minimizing the said peak varies from May to August 2023.

**CONCLUSIONS:** Our results indicate that if the epidemic continues, the best timing for vaccinations must be earlier than specified by the current plan in Korea. Further monitoring of outbreak trends is crucial for determining the optimal timing of vaccinations to manage future surges.

## Introduction

Since the Omicron wave in early 2022, the Korean government has been continuously lifting non-pharmaceutical interventions (NPIs), with the social distancing strategy having officially been lifted on April 18 of the same year.^1^ Furthermore, the obligation to wear masks in outdoor settings was changed to a recommendation on September 26, 2022, and the indoor mask-wearing policy was relaxed on January 1, 2023 with the exceptions of public transportation and hospitals. The public transportation requirement was subsequently relaxed as well on March 20, 2023.^2,3,4^ Consequently, the only currently active NPI in Korea is the mask mandate for hospitals. As of May 2023, approximately 20,000 cases are confirmed and 200 severely ill patients are hospitalized on a daily basis.^5^ The search for a maximally effective vaccination strategy is critical because COVID-19 must ideally be managed as a respiratory tract infection using medical tools without reapplication of NPIs unless more virulent variants emerge in the future.

Because mathematical modeling is a useful tool when devising intervention strategies and predictions, mathematical models have been designed to provide scientific insight into vaccination strategies. Research on prioritization according to age group, region, and risk level in relation to vaccination has been conducted since early 2021, when vaccinations began. Ko et al. developed a mathematical model that considers age groups, and discussed a vaccine prioritization strategy (by age) depending on the outbreak situation.^6^ Matrajt et al. suggested that vaccinations must be prioritized for individuals who are at high risk of severe disease or death.^7^ Similarly, Bubar et al. evaluated the effectiveness of different vaccine prioritization strategies by considering serostatus.^8^ Moore et al. found that combining vaccination with NPIs was the most effective strategy for controlling the spread of COVID-19.^9^ Following the 2023 Omicron wave, Elisha et al. discussed how waning immunity and the emergence of new variants will shape the long-term burden and dynamics of COVID-19 under consideration of various factors.^10^ However, their model fails to consider residual immunity against the severity induced by natural infection.

The Korean government changed the vaccination schedule to an annual basis from March 22, 2023, and announced that vaccinations will initiate between October and November of 2023.^11^ Immunity induced by vaccines and infections can wane over time, leading to an epidemic wave.^12^ The question of “To whom and when would be the best vaccination strategy?” is an issue that is currently being raised, as is the start of future surges. In this paper, we discuss effective vaccination strategies for managing COVID-19 surges using an advanced mathematical model. Based on open data and a national antibody survey, our model generates a detailed level of immunity according to whether an individual has been vaccinated or infected.

## Methods

### Mathematical modeling of COVID-19 considering immunity induced by vaccine or infection

In this study, the entire population wa divided into five groups:0-19 years old (*Ⅰ*), 20-49 years old (*Ⅱ*), 50-64 years old (*Ⅲ*), 65 years and older (*Ⅳ*), and medical personnel (*Ⅴ*). Individuals within these groups were classified further to reflect vaccination status as follows: unvaccinated, received primary vaccination less than six months ago, received primary vaccination six or more months ago, received booster vaccination (including third and three doses) less than six months ago, received booster vaccination (including bivariant vaccine) six or more months ago, and received the bivariant vaccine less than six months ago. Finally, they were categorized further with respect to prior infection status. Our model is represented as a flowchart (**Figure 1**) that considers population-level immunity, unreported cases, and patients in severe stages. Note that the subscript (*x*) of classes (*S*, *E*, *I*, *C*, *M*, *R*, 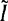, *D*) indicates the immunity-related status of the hosts, and the superscript (*i*) indicates the age group.

**Figure 1.**
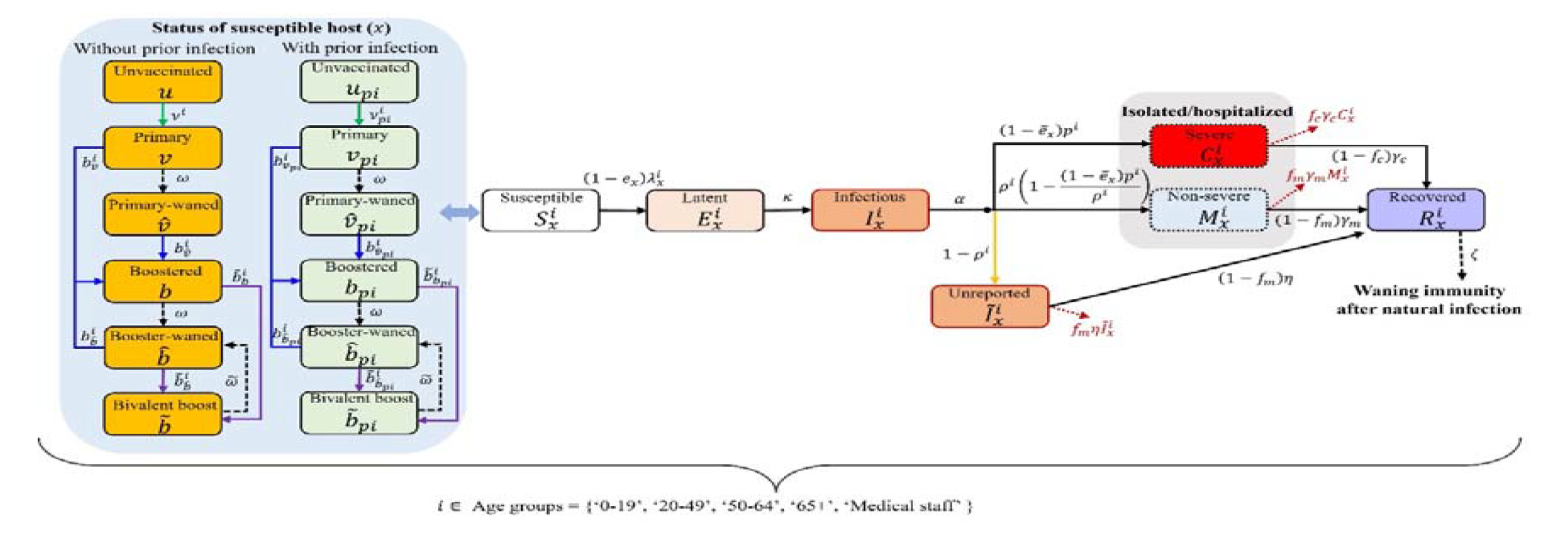
Flow chart for mathematical modeling of COVID-19

The transmission matrix between age groups encompasses households, school, work, hospitals, and other venues.^13, 14^ Statistics of outpatient/inpatient patients by age group, as well as the assumed numbers of contacts for outpatients (one for doctors and one for nurses) and inpatients (two for doctors and three for nurses) are also reflected in the matrix. Assuming that the contact and transmission patterns in work, school, and other venues have seasonality, the generalized formula for elements that constitute the transmission matrix is expressed as follows:

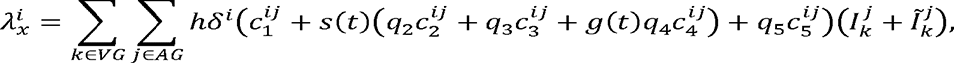

where *h* is the probability of infection through household contact; *δ^i^* is the age-dependent relative susceptibility compared to age 0-19 (i.e., δ*^l^* = 1) considering clinical susceptibility and other factors, such as compliance with policy or behavior; *q*_2_, *q*_3_, *q*_4_, and *q*_5_ are relative risks of infection through contacts at work, other venues, school, and hospital, respectively. We assume the risk of infection through contact in school to be the same as that in households – i.e., *q*_4_ = 1 – but affected by the seasonal factor (*s*(*t*) which is estimated every four weeks, and the school operation factor (*g*(*t*), which is 0 during vacation periods and 1 otherwise. In other words, contact in school is assumed to be absent during vacation periods. The relative risks associated with work-related and miscellaneous contacts – *q*_2_ and *q*_3_ – were both set to 0·30, and *q*_5_ was set to 0·06, reflecting the effectiveness of hospital mask policies.^15,16,17^ Note that force of infection is reduced by (1−*e_x_*) if the susceptible host has been vaccinated and/or infected previously. Similarly, the corresponding severity rate is reduced by 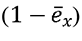. The unknown parameters *s*(*t*), *h*, and *δ^i^* are estimated using the cumulative number of confirmed cases by age. Detailed explanations of the formulae, estimated parameters, and results can be found in the **Supplementary material**. In this study, the difference between the proportion of confirmed cases and the positive rate of N-antibodies within each age group was reflected in the model as an unreported rate (1- *ρ_i_*).

To set the initial conditions for the mathematical model, we considered the groups with and without prior infections by vaccination history based on vaccination statistics and the results of the second national antibody survey, which contained the N-antibody positive rate, conducted in mid-December.^18^ In this process, the population with prior infection was adjusted considering the effects of vaccination. Consider a situation where vaccine effectiveness is 50%, and there are 100 vaccinated and 50 unvaccinated individuals for every 100 individuals with prior infection. The number of vaccinated people with prior infection is then 25 (50 × 100 × 0 · 5÷(100 × 0 · 5 + 50) = 25), whereas the number of unvaccinated people with prior infection is 25 (50 × 50÷(100 × 0 · 5+50)= 25). In this study, half of the previously infected population was assumed to belong to 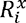, which still had full immunity.

### Vaccination-time-dependent forecast scenario design

We conducted a simulation of COVID-19 outbreaks in Korea up to the current situation to perform parameter estimation. To derive the appropriate timing for vaccination, the simulation was subsequently extended for one year beginning with May 11, 2023. To set the vaccination amount, we referred to the number of doses administered during the winter vaccination period at the end of 2022 (approximately 6·6 million doses per 100 days).^19^ We fixed that 5 million doses of the bivalent vaccine were administered for 100 days, which is less than the results of the last winter vaccination period. Scenarios vary by setting different vaccination target groups, extra doses, and outbreak trends. The timing of vaccination was set from the model simulation extension start date (May 11, 2023) to 300 days later (March 6, 2024) at intervals of 10 days. During the extension period, we set up antiviral medicines, and 40% of the patients over 50 years of age were prescribed the default settings. The use of antiviral drugs yielded a 51% reduction in the number of severe cases.^20^

## Results

The values and distributions of the estimated parameters are included in **Supplementary material**; the seasonal factor *s*(*t*) was estimated to cyclically change with 0·49, 0·19, 0·82, 0·50, and 0·57 at four-week intervals. The model simulation results of the data fitting and one-year extension are displayed in **Figure 2**. As an extension, the seasonal factor in the last phase (from April 6 to May 11, 2023) was used, and vaccinations and antiviral medicines were not administered. The next peak is expected to occur in mid-October 2023. Note that the sudden decrease in the underage group in July was due to summer vacation.

**Figure 2.**
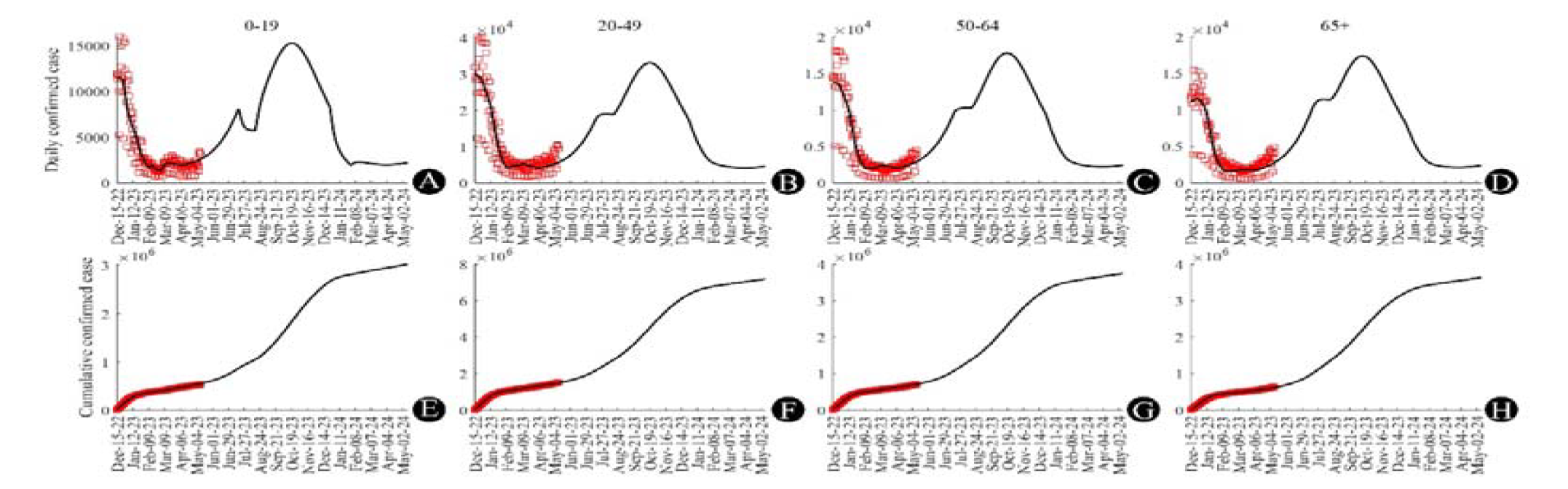
Parameter estimation and one-year extension simulation results. (A) to (D) ((E) to (F)) represent daily confirmed cases (cumulative confirmed cases) within each age group. Dark curves and red squares denote model simulation results and real-world data, respectively.

The simulation results reflecting different vaccination target settings are presented in **Figure 3**. If a vaccination program is implemented earlier, the peak will be delayed from mid-October to mid-November 2023. The daily and cumulative numbers of cases were lower when all adults were the targets of vaccination, whereas the number of severe cases was lower when elderly people were vaccinated. **Figure 4** summarizes the outbreak outcome forecasts based upon initial vaccination timing. If vaccination is initiated at the end of May targeting individuals older than 65 (wherein approximately 30,000 daily confirmed cases occur), the peak number of hospitalized severe patients could be minimized to 480. **Table 1** summarizes the vaccination schedules expected to minimize the cumulative or peak number of cases (or hospitalized patients with severe symptoms), as well as predicted outbreak outcomes depending on different scenario characteristics.

**Figure 3.**
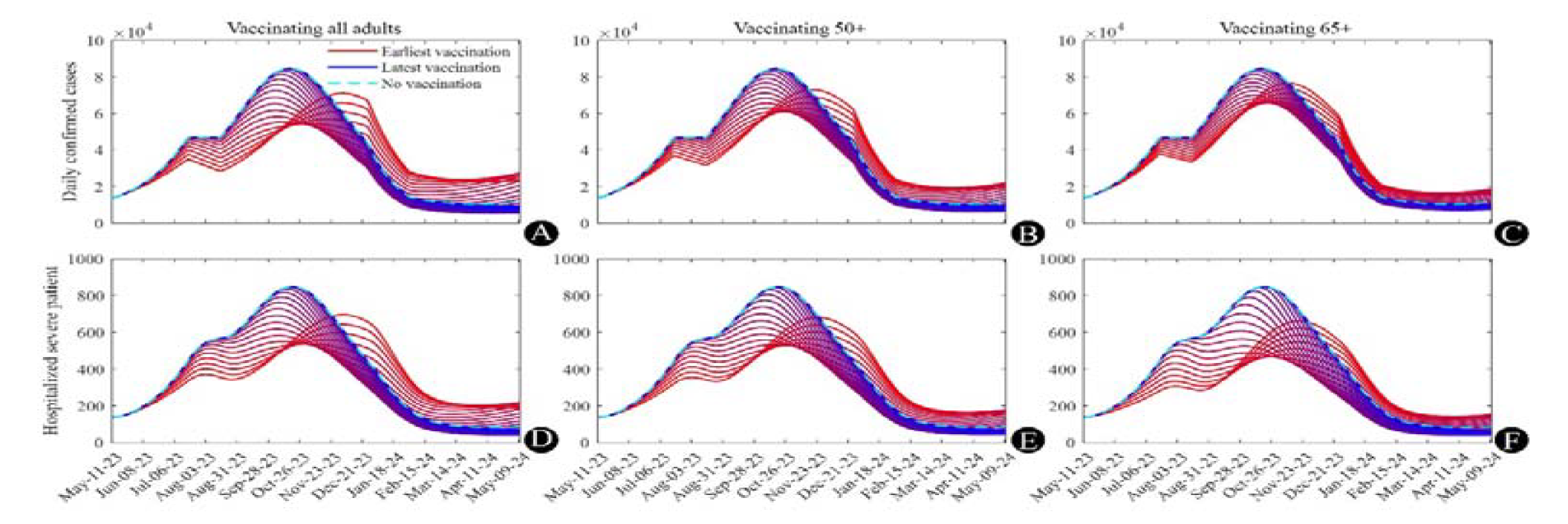
Time series extension simulation results, (A) to (C) represent daily confirmed cases and (D) to (F) measure hospitalizations with severe symptoms, with different vaccination target settings (each column) and times of vaccination. Dashed cyan curves indicate simulation results without vaccination, and all other curves represent simulation results under various vaccination conditions. Curves are depicted in a gradient between red and blue in ascending order of first vaccination time.

**Figure 4.**
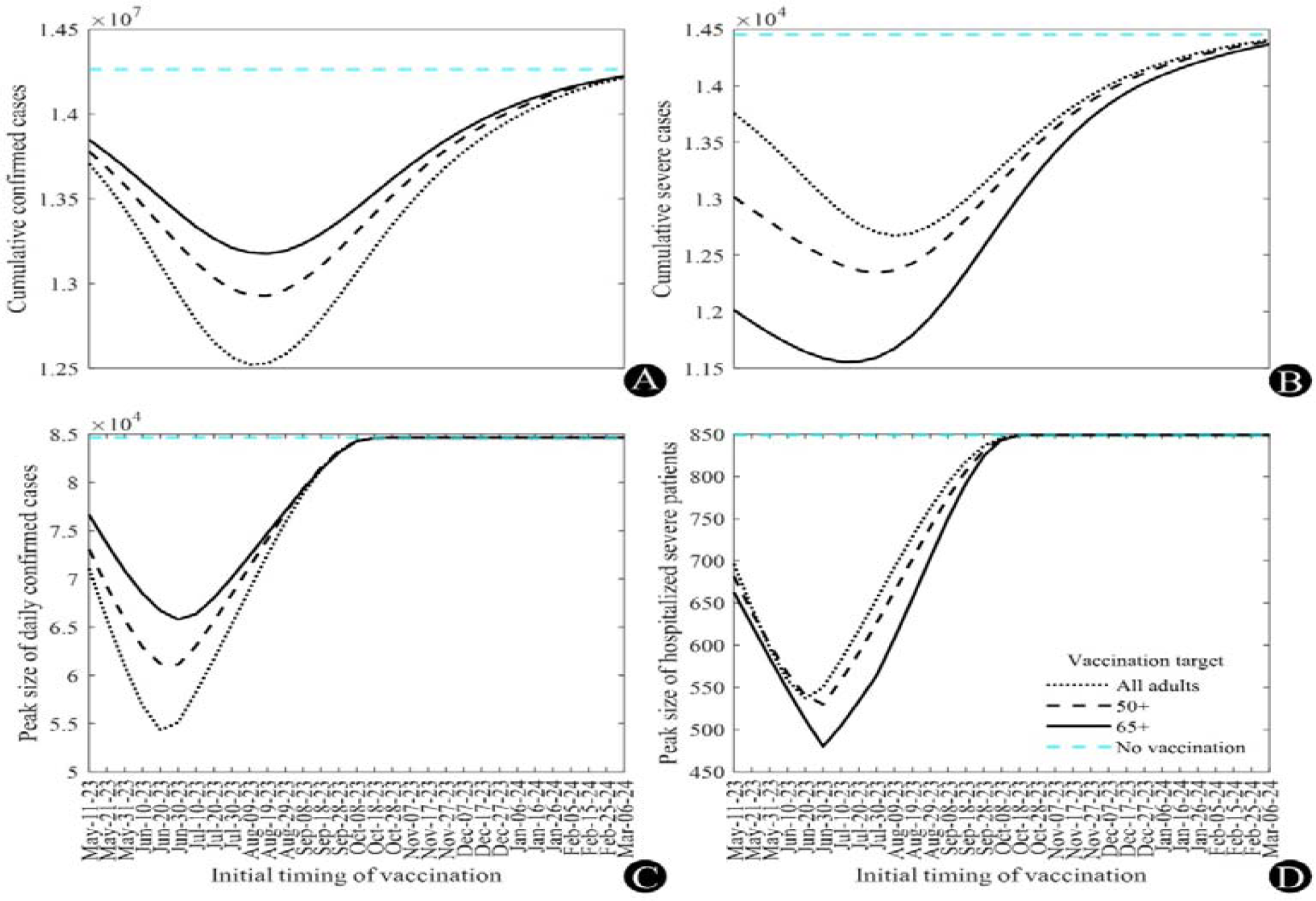
Extended simulation results from different vaccination targets and initial times of vaccination. (A) and (B) show the number of cumulative confirmed and severe cases, whereas (C) and (D) depict the peaks of daily confirmed cases and hospitalizations with severe conditions, respectively. Note that the x-axis indicates the initial timing of vaccination.

**Table 1.** Lists of the best vaccination timing and outbreak outcomes according to the extended simulation setting

| Number of doses and vaccination target age |  | 5 million,<br>All adults | 5 million,<br>Age over 50 | 5 million,<br>Age over 65 | 2.5 million,<br>Age over 65 | 7.5 million,<br>Age over 65 | 7.5 million,<br>5 million<br>for age over<br>65 and 2.5<br>for others | 10 million,<br>5 million<br>for age over<br>65 and 5 for<br>others | 5 million,<br>Age over 65 | 5 million,<br>Age over 65 |
| --- | --- | --- | --- | --- | --- | --- | --- | --- | --- | --- |
| Outbreak trend |  | Current | Current | Current | Current | Current | Current | Current | -20% | +20% |
| Cumulative<br>case<br>minimization | Best timing | August 9 | August 9 | August 19 | September 8 | July 30 | August 9 | August 9 | September 8 | August 9 |
|  | Case number<br>(million) | 12.52 | 12.93 | 13.18 | 13.66 | 12.86 | 12.25 | 11.26 | 10.70 | 14.98 |
| Cumulative<br>severe case<br>minimization | Best timing | August 9 | July 30 | July 10 | August 9 | June 20 | July 30 | July 30 | August 29 | May 31 |
|  | Case number<br>(thousand) | 12.67 | 12.35 | 11.56 | 12.89 | 10.47 | 11.00 | 10.36 | 10.27 | 12.23 |
| Peak case<br>size<br>minimization | Best timing | June 20 | June 30 | June 30 | July 30 | June 10 | June 30 | June 30 | August 19 | May 21 |
|  | Peak size<br>(thousand) | 54.37 | 61.12 | 65.80 | 72.98 | 64.20 | 50.70 | 42.27 | 50.21 | 81.84 |
| Peak size of<br>administered<br>severe case<br>minimization | Best timing | June 20 | June 30 | June 30 | August 9 | May 31 | June 20 | June 20 | August 19 | May 11 |
|  | Peak size | 537 | 530 | 480 | 610 | 420 | 430 | 406 | 401 | 618 |

Because we found that vaccination schemes targeting individuals older than 65 minimize the number of severe cases, we performed an additional scenario-based study where we fixed the number of doses for the over-65 age group at 5 million and retained the current outbreak trend, but varied additional doses for other adult groups between 2·5 and 5 million (**Figure 5**). As the number of extra doses increased, both the cumulative and peak numbers of severe patients were minimized if vaccination initiated at the optimal time. However, the additional benefits decreased if vaccination initiated earlier or later than the appropriate timing.

**Figure 5.**
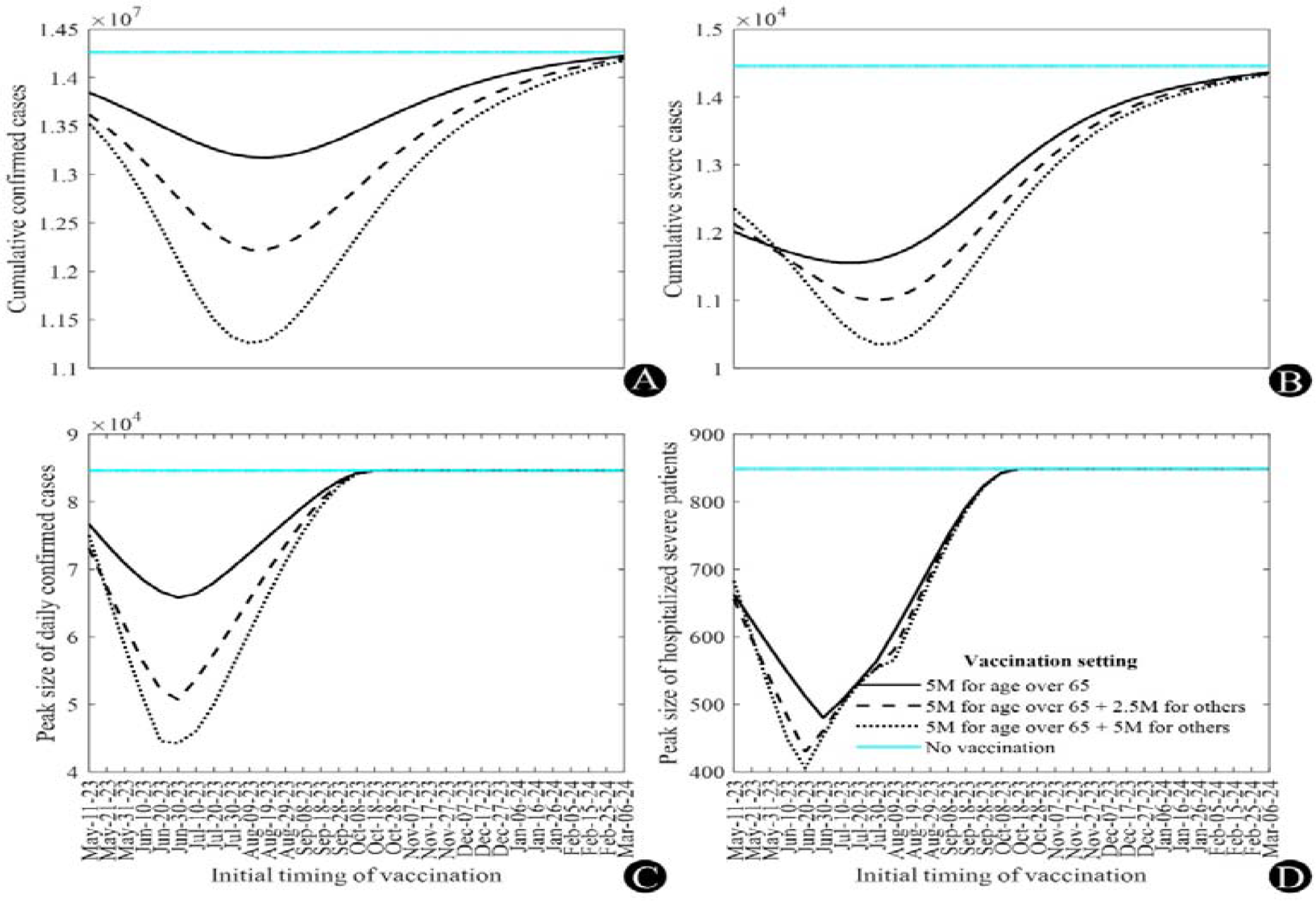
Extended simulation results with different vaccine dosage amounts and initial times of vaccination. (A) and (B) measure cumulative confirmed and severe cases, whereas (C) and (D) depict the peaks of daily confirmed cases and severe hospitalizations, respectively. Note that the x-axis indicates initial time of vaccination.

When additional analyses were performed for scenarios in which different numbers of individuals older than 65 were vaccinated, the optimal timing of vaccination differed accordingly (August 9 for 2.5 million and May 31 for 7.5 million), even assuming the same outbreak trend (**Table 1**).

Here, we fixed the vaccination setting at 5 million for those aged > 65 years (solid curves in **Figure 6**) and extended the simulation results to different outbreak trends (*s*(*t*) in the model). The best time to minimize the cumulative or peak numbers of cases and severe hospitalizations can be delayed if s(t) decreases; however, vaccination should initiate earlier if s(t) increases. If *s*(*t*) decreases by 20%, the best initial time of vaccination to minimize the peak number of severe patients is delayed from June 30 to August 19, 2023.

**Figure 6.**
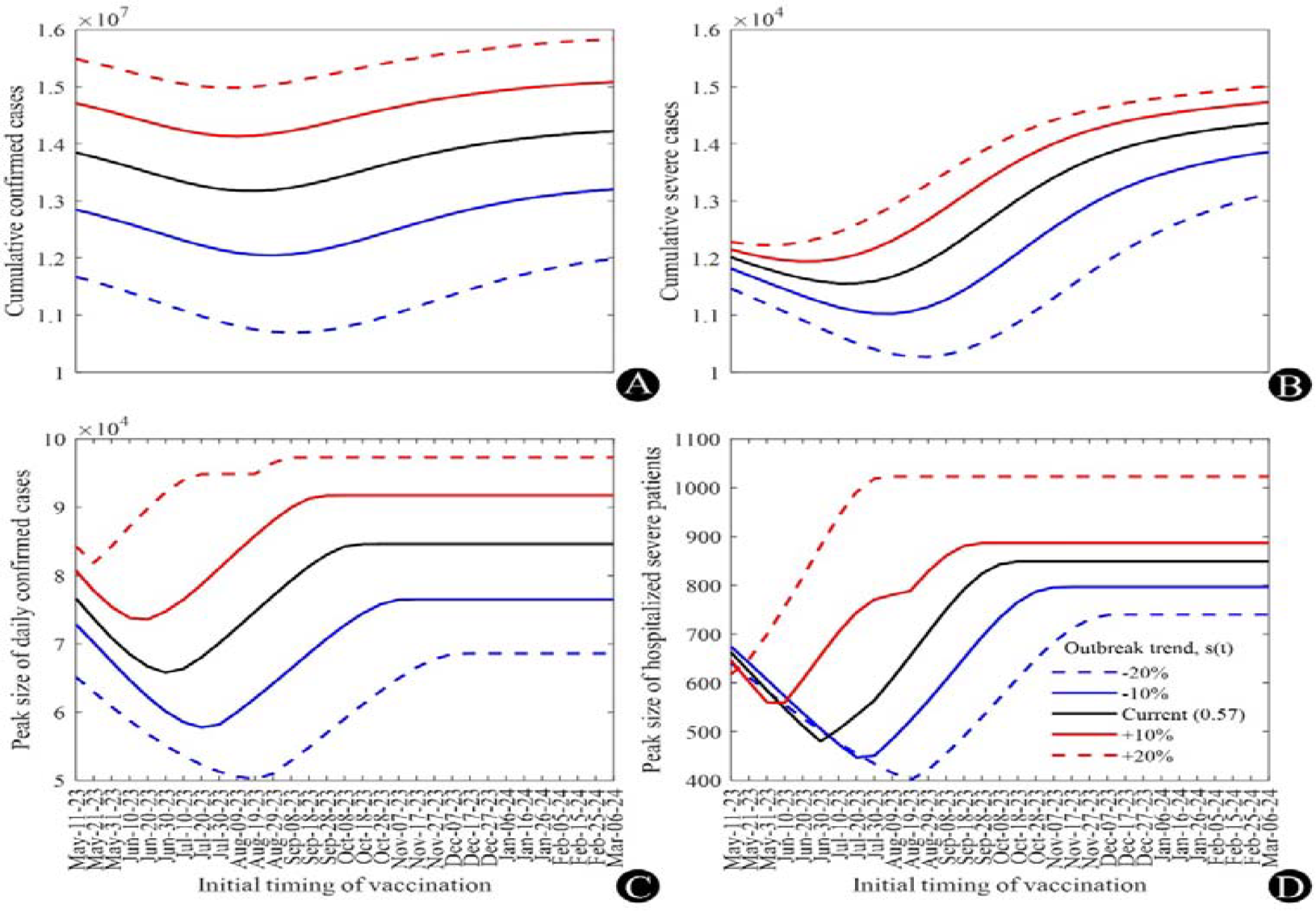
Extended simulation results with different vaccination outbreak trends and times of vaccination. (A) and (B) measure cumulative confirmed and severe cases, whereas (C) and (D) represent peaks of daily confirmed cases and severe hospitalizations, respectively. Note that the x-axis indicates the initial time of vaccination.

## Discussion

As the mathematical model increases in complexity, the required data features become more challenging to aggregate. Furthermore, if important parameters are assumed or estimated, overfitting may lead to incorrect decisions or predictions^21, 22^ In this study, we developed an advanced model with precisely set data-based parameters and initial conditions. We considered hosts who experienced infection before the initial time, and the antibody survey may help set the initial conditions of the system ODEs.^18^

As of May 11, 2023, if the current trend is extended without additional vaccination, the peak of the next wave is predicted to occur in mid-October, and the number of severe hospitalized cases may exceed 800. Currently, the number of intensive care unit beds for COVID-19 patients in Korea is 249, which is below the expected peak size.^23^ Our results emphasize the need to securely prepare medical resources to respond to the next wave, and further suggest that that the next wave can be managed without the use of NPIs.

Our simulation results suggest that the optimal timing of vaccination is earlier than that specified in the current governmental plan, as we found that the peak number of severe hospitalizations was minimized with an initial vaccination time ranging from May to August depending on the outbreak trend.^11^ In the extended simulation results using the current outbreak trend, we found the initial vaccination time to minimize the said peak to occur at the end of June. On the same day, the number of daily confirmed cases is projected to reach approximately 30,000, indicating the early phase of resurgence. However, it is challenging to determine the best timing for vaccination given an increasing number of cases. Therefore, it is necessary to compare future trends with model simulation results to determine effective vaccination scheduling. By reflecting varying outbreak trends, our model would help determine the best timing for vaccine administration.

It is apparent that as the scale of vaccination increases, the outbreak scale decreases. However, we observed that rushed large-scale vaccination did not have a significant effect compared to a scenario with lower doses (refer to panels (B) and (D) in **Figure 5**.) because the immunity induced by vaccines wanes faster than that induced by natural infections. Similarly, we observed the best timing to change with respect to vaccine dosage (**Table 1**) given a fixed outbreak trend. Therefore, the timing of vaccination must be optimized not only by the outbreak trend, but also by the expected vaccination scale.

The limitations of this study were as follows. First, the contact matrix was set based on data preceding the COVID-19 pandemic, and may differ due to changes in population behaviors or lifestyle patterns following the outbreak. Second, although the proportion of the population reporting or self-isolating may have changed after December 2022, this information is reflected in the model with the same values as before. Finally, we did not consider new variants or lifted policies that may change outbreak trends. The actual progression of resurgence would differ from the prediction due to changes in control strategies for COVID-19 implemented since early June this year.

## Conclusion

The effectiveness of vaccinations decreases over time, raising concerns about vaccination timing in response to resurgence. In this study, we simulated the immune status of a population with a model that considers antibody survey data, and determined the optimal vaccination strategy to control the endemic population. We conducted an analysis of the appropriate timing of vaccination, and found that if the current trend (as of May 11, 2023) was to continue, the vaccination timing required to minimize the peak number of hospitalized patients would range from mid-May to mid-August, depending on the vaccination scale and outbreak trend. We expect that our simulation results will help health authorities determine appropriate vaccination scheduling procedures.

## Data Availability

All data produced in the present study are available upon reasonable request to the authors

## Acknowledgments

This paper is supported by the Korea National Research Foundation (NRF) grant funded by the Korean government (MEST) (NRF-2021M3E5E308120711). This paper is also supported by the Korea National Research Foundation (NRF) grant funded by the Korean government (MEST) (NRF- 2021R1A2C100448711). This study was conducted utilizing the database of the 2023 Community-Based Representative Sample COVID-19 Seroprevalence Survey by the Korea National Institute of Health.

## References

1 Back to normal life, complete lifting of social distancing measures from April 18. Seoul Metropolitan Government. Available from: https://news.seoul.go.kr/welfare/archives/542308 (Korean, author’s translation)

2 Lifting of mandatory outdoor mask wearing and recommendation of voluntary wearing. Korea Disease Control and Prevention Agency. Available from: https://www.kdca.go.kr/gallery.es?mid=a20503010000&bid=0002&list_no=145857&act=view (Korean, author’s translation)

3 Where should I wear a mask indoors?. Korea Disease Control and Prevention Agency. Available from: https://www.kdca.go.kr/board/board.es?mid=a20501050000&bid=0015&act=view&list_no=721800 (Korean, author’s translation)

4 Mandatory mask wearing on public transportation will be lifted from the April 20. Korea Policy Briefing. Available from: https://www.korea.kr/multi/visualNewsView.do?newsId=148912938 (Korean, author’s translation)

5 Domestic occurrence status of COVID-19. Korea Disease Control and Prevention Agency. Available from: https://ncov.kdca.go.kr/bdBoardList_Real.do?brdId=1&brdGubun=11&ncvContSeq=&contSeq=&board_id=&gubun<u>=</u> (Korean, author’s translation)

6 Ko Y, Lee J, Seo Y, Jung E. Risk of COVID-19 transmission in heterogeneous age groups and effective vaccination strategy in Korea: a mathematical modeling study. Epidemiology and Health. 2021;43.

7 Matrajt L, Eaton J, Leung T, Brown ER. Vaccine optimization for COVID-19: Who to vaccinate first?. Science Advances. 2021 Feb 3;7(6):eabf1374.

8 Bubar KM, Reinholt K, Kissler SM, Lipsitch M, Cobey S, Grad YH, Larremore DB. Model-informed COVID- 19 vaccine prioritization strategies by age and serostatus. Science. 2021;371(6532):916–21.

9 Moore S, Hill EM, Tildesley MJ, Dyson L, Keeling MJ. Vaccination and non-pharmaceutical interventions for COVID-19: a mathematical modelling study. The lancet infectious diseases. 2021;21(6):793–802.

10 Are EB, Song Y, Stockdale JE, Tupper P, Colijn C. COVID-19 endgame: From pandemic to endemic? Vaccination, reopening and evolution in low-and high-vaccinated populations. Journal of Theoretical Biology. 2023;559:111368.

11 COVID-19 vaccine, switching to once a year vaccination. Korea Disease Control and Prevention Agency. Available from: https://ncov.kdca.go.kr/tcmBoardView.do?gubun=BDJ&brdId=3&brdGubun=31&dataGubun=&ncvContSeq=7216&board_id=312&contSeq=7216 (Korean, author’s translation)

12 Goldberg Y, Mandel M, Bar-On YM, Bodenheimer O, Freedman L, Haas EJ, et al. Waning immunity after the BNT162b2 vaccine in Israel. New England Journal of Medicine. 2021;385(24):e85.

13 Prem K, Cook AR, Jit M. Projecting social contact matrices in 152 countries using contact surveys and demographic data. PLoS computational biology. 2017;13(9):e1005697.

14 National health care statistics: number of inpatients by patient residence, gender, and age. Ministry of Health and Welfare of Korea; Available from: https://kosis.kr/statHtml/statHtml.do?orgId=117&tblId=DT_117049_A055&conn_path=I2 (Korean, author’s translation)

15 Zee ST, Kwok LF, Kee KM, Fung LH, Luk WP, Chan TL, et al. Impact of COVID-19 Vaccination on Healthcare Worker Infection Rate and Outcome during SARS-CoV-2 Omicron Variant Outbreak in Hong Kong. Vaccines. 2022;10(8):1322.

16 Cheng H, Jian S, Liu D, et al. Contact Tracing Assessment of COVID-19 Transmission Dynamics in Taiwan and Risk at Different Exposure Periods Before and After Symptom Onset. JAMA Intern Med. 2020;180(9):1156–1163.

17 Payne DC, Smith-Jeffcoat SE, Nowak G, Chukwuma U, Geibe JR, Hawkins RJ, et al. SARS-CoV-2 infections and serologic responses from a sample of US Navy service members—USS Theodore Roosevelt, April 2020. Morbidity and Mortality Weekly Report. 2020;69(23):714.

18 Announcement of the results of the second survey on the positive rate of COVID-19 antibodies, Korea Disease Control and Prevention Agency. Available from: https://www.kdca.go.kr/gallery.es?mid=a20503010000&bid=0002&list_no=145991&act=view (Korean, author’s translation)

19 Daily vaccination status. Korea Disease Control and Prevention Agency. Available from: https://ncv.kdca.go.kr/vaccineStatus.es?mid=a11710000000 (Korean, author’s translation)

20 Jeon J, Chin B. Treatment Options for Patients With Mild-to-Moderate Coronavirus Disease 2019 in Korea. Journal of Korean Medical Science. 2022;37(48).

21 Basu S, Andrews J. Complexity in mathematical models of public health policies: a guide for consumers of models. PLoS medicine. 2013;10(10):e1001540.

22 James LP, Salomon JA, Buckee CO, Menzies NA. The use and misuse of mathematical modeling for infectious disease policymaking: lessons for the COVID-19 pandemic. Medical Decision Making. 2021;41(4):379–85.

23 Overall status of COVID-19 in Korea. Korea Disease Control and Prevention Agency. Available from: https://ncov.kdca.go.kr/ (Korean, author’s translation)

