## supplementary material for "Effective vaccination strategies to control COVID-19 outbreak: A modeling study"

**System equations of the mathematical model of COVID-19 outbreaks in Korea**

The system of equations describing our model is as follows,

$$\frac{dS_{x}^{i}}{dt}=-\lambda_{x}^{i}S_{x}^{i}+IN_{x}^{i}-OUT_{x}^{i},$$

$$\frac{dE_{x}^{i}}{dt}=\lambda_{x}^{i}S_{x}^{i}-\kappa E_{x}^{i},$$

$$\frac{dI_{x}^{i}}{dt}=\kappa E_{x}^{i}-\alpha I_{x}^{i},$$

$$\frac{dM_{x}^{i}}{dt}=\alpha\rho^{i}\left( 1-\frac{\left( 1-\bar{e}_{x} \right)p^{i}}{\rho^{i}} \right)I_{x}^{i}-\gamma_{m}M_{x}^{i},$$

$$\frac{dC_{x}^{i}}{dt}=\alpha\left( 1-\bar{e}_{x} \right)p^{i}I_{x}^{i}-\gamma_{c}C_{x}^{i},$$

$$\frac{d\tilde{I}_{x}^{i}}{dt}=\alpha\left( 1-\rho^{i} \right)I_{x}^{i}-\eta\tilde{I}_{x}^{i},$$

$$\frac{dR_{x}^{i}}{dt}=\left( 1-f_{m} \right)\gamma_{m}M_{x}^{i}+\left( 1-f_{c} \right)\gamma_{c}C_{x}^{i}+\left( 1-f_{m} \right)\eta\tilde{I}_{x}^{i}-\zeta,$$

$$\frac{dD_{x}^{i}}{dt}=f_{m}\gamma_{m}M_{x}^{i}+f_{c}\gamma_{c}C_{x}^{i}+f_{m}\eta\tilde{I}_{x}^{i},$$

$$\lambda_{x}^{i}=\sum_{k\in VG} \sum_{j\in AG} h\left( c_{1}^{ij}+s\left( t \right)\left( q_{2}c_{2}^{ij}+q_{3}c_{3}^{ij}+g\left( t \right)q_{4}c_{4}^{ij} \right)+q_{5}c_{5}^{ij} \right)\left( {I_{k}^{j}+\tilde{I}}_{k}^{j} \right),$$

$$IN_{u}^{i}=0, {OUT}_{u}^{i}=\nu^{i},$$

$$IN_{v}^{i}=\nu^{i}, {OUT}_{v}^{i}=\omega S_{v}^{i}+b_{v}^{i},$$

$$IN_{\hat{v}}^{i}=\omega S_{v}^{i}, {OUT}_{\hat{v}}^{i}=b_{\hat{v}}^{i},$$

$$IN_{b}^{i}=b_{v}^{i}+b_{\hat{v}}^{i}+b_{\hat{b}}^{i}, {OUT}_{b}^{i}=\omega S_{b}^{i}+\tilde{b}_{b}^{i},$$

$$IN_{\hat{b}}^{i}=\omega S_{b}^{i}+\tilde{\omega}S_{\tilde{b}}^{i}, {OUT}_{\hat{b}}^{i}=b_{\hat{b}}^{i}+\tilde{b}_{\hat{b}}^{i},$$

$$IN_{\tilde{b}}^{i}=\tilde{b}_{b}^{i}+\tilde{b}_{\hat{b}}^{i}, {OUT}_{\tilde{b}}^{i}=\tilde{\omega}S_{\tilde{b}}^{i},$$

$$IN_{u_{pi}}^{i}=\zeta\left( R_{u}^{i}+R_{u_{pi}}^{i} \right), {OUT}_{u_{pi}}^{i}=\nu_{pi}^{i},$$

$$IN_{v_{pi}}^{i}=\nu_{pi}^{i}, {OUT}_{v_{pi}}^{i}=\omega S_{v_{pi}}^{i}+b_{v_{pi}}^{i},$$

$$IN_{\hat{v}_{pi}}^{i}=\zeta\left( R_{v}^{i}+R_{\hat{v}}^{i}+R_{v_{pi}}^{i}+R_{\hat{v}_{pi}}^{i} \right)+\omega S_{v_{pi}}^{i}, {OUT}_{\hat{v}_{pi}}^{i}=b_{\hat{v}_{pi}}^{i},$$

$$IN_{b_{pi}}^{i}=b_{v_{pi}}^{i}+b_{\hat{v}_{pi}}^{i}+b_{\hat{b}_{pi}}^{i}, {OUT}_{b_{pi}}^{i}=\omega S_{b_{pi}}^{i}+\tilde{b}_{b_{pi}}^{i},$$

$$IN_{\hat{b}_{pi}}^{i}=\zeta\left( R_{b}^{i}+R_{\hat{b}}^{i}+R_{\tilde{b}}^{i}+R_{b_{pi}}^{i}+R_{\hat{b}_{pi}}^{i}+R_{\tilde{b}_{pi}}^{i} \right)+\omega S_{b_{pi}}^{i}+\tilde{\omega}S_{\tilde{b}_{pi}}^{i}, {OUT}_{\hat{b}_{pi}}^{i}=b_{\hat{b}_{pi}}^{i}+\tilde{b}_{\hat{b}_{pi}}^{i},$$

$$IN_{\tilde{b}_{pi}}^{i}=\tilde{b}_{b_{pi}}^{i}+\tilde{b}_{\hat{b}_{pi}}^{i}, {OUT}_{\tilde{b}_{pi}}^{i}=\tilde{\omega}S_{\tilde{b}_{pi}}^{i},$$

where $x∊VG=\{u,v,\hat{v},b,\hat{b},\tilde{b},u_{pi},v_{pi},\hat{v}_{pi},b_{pi},\hat{b}_{pi},\tilde{b}_{pi}\}$, $i∊AG=\{Ⅰ,Ⅱ,Ⅲ,Ⅳ,Ⅴ\}$, and vaccine administration per day parameters, {$\nu^{i}$, $b_{v}^{i}$, $b_{\hat{v}}^{i}$, $b_{\hat{b}}^{i}$, $\tilde{b}_{b}^{i}$, $\tilde{b}_{\hat{b}}^{i}$, $\nu_{pi}^{i}$, $b_{v_{pi}}^{i}$, $b_{\hat{v}_{pi}}^{i}$, $b_{\hat{b}_{pi}}^{i}$, $\tilde{b}_{b_{pi}}^{i}$, $\tilde{b}_{\hat{b}_{pi}}^{i}$}, correspond to the data on daily number of administered vaccines.^[1],^^[2]^ Description and value of model parameters are listed in **Supplementary Table 1**. In our model, two types of vaccine effectiveness are used: against infection ($e_{x}$) and against severity ($\bar{e}_{x}$). The values were adjusted according to the effectiveness and proportion of an age group vaccinated with the different types of vaccines administered in Korea. The details of the calculation are described in our past research.^[3]^ For hosts with hybrid immunity (prior infection + vaccination), we assumed that their waned-vaccine effectiveness against infection is the average value between the unvaccinated but infected ($e_{u_{pi}}$,) and vaccinated ($e_{v_{pi}}$ or $e_{b_{pi}}$ or $e_{\tilde{b}_{pi}}$). For bivariant vaccines, we assumed that the effectiveness against infection and severity are 0·8 and 0·95 (0·8 and 0·95), respectively, if the host does not have a prior infection (host has prior infection). **Supplementary Table 2.** shows the values for the vaccine effectiveness that were used in this study.

**Supplementary Table 1.** Model parameters

| **Symbol** | **Description** | **Value** | **Reference** |
| --- | --- | --- | --- |
| $q_{2}$ | Relative risk of work contact compared to household contact | 0·3035 | ^[4]^ |
| $q_{3}$ | Relative risk of other contact compared to household contact | 0·3035 | ^4^ |
| $q_{4}$ | Relative risk of school contact compared to household contact | 1 | Assumed |
| $q_{5}$ | Relative risk of hospital contact compared to household contact | 0·057 | ^[5],^^[6]^ |
| $1/\kappa$ | Latent period | 2 (days) | ^[7],^^[8],^^[9],^^[10]^ |
| $1/\alpha$ | Infectious period of reported case | 4 (days) | ^10,^^[11]^ |
| $1/\eta$ | Extra infectious period of unreported case | 2 (days) | ^[12]^ |
| $\rho^{i}$ | Report rate | 91·91% (age 0-19)  80·87% (age 20-49)  64·51% (age 50-64)  68·85% (age 65+)  95·00% (MS) | Assumed: MS |
| $p^{i}$ | Case severe rate | 0·01% (age 0-19)  0·10% (age 20-29)  0·37% (age 30-49)  0·95% (age 50-64)  0·10% (MS) | ^[13],^^[14]^ |
| $\gamma_{c}$ | Recovery period of severe case | 10 (days) | ^[15]^ |
| $\gamma_{m}$ | Recovery period of mild case | 7 (days) | ^[16]^ |
| $f_{c}$ | Case fatality rate of severe case | 28·55% | ^[17]^ |
| $f_{m}$ | Case fatality rate of mild case | 0·0070% | ^13^ |
| $1/\omega$ | Waning period of vaccine-induced immunity | 180 (days) | ^[18]^ |
| $1/\tilde{\omega}$ | Waning period of bivariant vaccine-induced immunity | 180 (days) | Assumed |
| $1/\zeta$ | Waning period of infection-induced immunity | 360 (days) | ^18^ |

*****Aggregated from the national seroprevalence survey

**Supplementary Table 2.** Vaccine effectiveness

| **VE against** | **Prior infection** | **Phase** | **Symbol** | **Value** | **Reference** |
| --- | --- | --- | --- | --- | --- |
| Infection | No | Primary | $e_{v}$ | 0·37 | ^[19],^^[20]^ |
|  |  | Primary-waned | $e_{\hat{v}}$ | 0·12 | ^19,20^ |
|  |  | Boosted | $e_{b}$ | 0·60 | ^19,20^ |
|  |  | Booster-waned | $e_{\hat{b}}$ | 0·13 | ^19,20^ |
|  |  | Bivalent boosted | $e_{\tilde{b}}$ | 0·80 | Assumed |
|  | Yes | Unvaccinated | $e_{u_{pi}}$ | 0·52 | ^18^ |
|  |  | Primary | $e_{v_{pi}}$ | 0·55 | ^18^ |
|  |  | Primary-waned | $e_{\hat{v}_{pi}}$ | 0·54 | Assumed |
|  |  | Boosted | $e_{b_{pi}}$ | 0·77 | ^18^ |
|  |  | Booster-waned | $e_{\hat{b}_{pi}}$ | 0·65 | Assumed |
|  |  | Bivalent boosted | $e_{\tilde{b}_{pi}}$ | 0·80 | Assumed |
| Severity | No | Primary | $\bar{e}_{v}$ | 0·77 | ^[21]^ |
|  |  | Primary-waned | $\bar{e}_{\hat{v}}$ | 0·55 | ^21^ |
|  |  | Boosted | $\bar{e}_{b}$ | 0·90 | ^21^ |
|  |  | Booster-waned | $\bar{e}_{\hat{b}}$ | 0·58 | ^21^ |
|  |  | Bivalent boosted | $\bar{e}_{\tilde{b}}$ | 0·95 | Assumed |
|  | Yes | Unvaccinated | $\bar{e}_{u_{pi}}$ | 0·75 | ^[22]^ |
|  |  | Primary | $\bar{e}_{v_{pi}}$ | 0·97 | ^22^ |
|  |  | Primary-waned | $\bar{e}_{\hat{v}_{pi}}$ | 0·97 | Assumed |
|  |  | Boosted | $\bar{e}_{b_{pi}}$ | 0·98 | ^22^ |
|  |  | Booster-waned | $\bar{e}_{\hat{b}_{pi}}$ | 0·97 | Assumed |
|  |  | Bivalent boosted | $\bar{e}_{\tilde{b}_{pi}}$ | 0·98 | Assumed |

**Initial condition estimation**

We divide the susceptible population in Korea considering variety of factors; Age, medical staff (MS), vaccine history, and the prior infection. Firstly, entire population is divided into 5 subgroups considering the host’s age and whether the host is medical staff or not; 0-19 ($Ⅰ$), 20-49 ($Ⅱ$), 50-64 ($Ⅲ$), 65+ ($Ⅳ$), and MS ($Ⅴ$).^[23],^^[24]^  Later, considering vaccination status in December 15, 2022, divided population is once more divided; unvaccinated, $u$, primary vaccinated within 180 days, $v$, primary vaccinated after 180 days, $\hat{v}$, boostered within 180 days, $b$, boostered after 180 days, $\hat{b}$, bivalent booster administrated, $\tilde{b}$. Considering high vaccination rate of MS, we set that 90% of MS are in boostered state (22·5% $b$, 45% $\hat{b}$, and 22·5% $\tilde{b}$) and 10% are in $u$. Still, we need to consider proportion of population who experienced infection. We adjusted the proportion considering population size, and set report rate as $\rho^{i}$, which is calculated using the difference between number of reported proportion and actual N-positive proportion. Since the investigation did not target age 0-4, we simply set that age 0-4 has same N-positive ratio with age 5-9. By using the N-positive ratio, age-vaccine-subgrouped population is once more subgrouped considering prior infection (subscript $pi$). In this case, vaccine effectiveness was considered to give more proportion of N-positive ratio to unvaccinated hosts. Since most of the COVID-19 infection in Korea occurred in 2022, we simply set that half of hosts with prior infection still remains in $R_{x}^{i}$. Initial state values are listed in **Supplementary Table 3**.

Since the outbreak situation in mid-August was relatively stable, we simply estimated the initial states of $E_{x}^{i}$, $I_{x}^{i}$, and $\tilde{I}_{x}^{i}$ considering endemic equilibrium. Administered severe patient data was not age-specified, but daily confirmed case data was. Therefore, we used confirmed case number by age data to estimate the initial states. Let [${DATA}_{i}$] is the age-specified data, then initial states are formulated as,

$$\sum_{k} I_{k}^{i}\left( 0 \right)=\frac{[DATA]}{\rho^{i}\alpha}$$

$$\sum_{k} E_{k}^{i}\left( 0 \right)=\frac{[DATA]}{\rho^{i}\kappa}$$

$$\sum_{k} \tilde{I}_{k}^{i}\left( 0 \right)=\frac{\left( 1-\rho^{i} \right)}{\rho^{i}}\frac{[DATA]}{\eta}.$$

(2)

**Supplementary Table 3.** Initial states of the susceptible groups

| **Prior infection** | **Age group** | **0-19** ($Ⅰ$) | **20-49** ($Ⅱ$) | **50-64** ($Ⅲ$) | **65+** ($Ⅳ$) | **MS** ($Ⅴ$) |
| --- | --- | --- | --- | --- | --- | --- |
| No | Unvaccinated | 676,846 | 302,637 | 207,094 | 6,381 | 0 |
|  | Primary | 13,133 | 9,752 | 2,300 | 3,072 | 0 |
|  | Primary-waned | 385,594 | 1,426,169 | 331,213 | 72,458 | 0 |
|  | Boosted | 30,925 | 190,466 | 1,200,399 | 823,945 | 13,376 |
|  | Booster-waned | 103,539 | 3,346,101 | 1,401,911 | 445,079 | 26,751 |
|  | Bivalent boosted | 30,547 | 594,641 | 1,158,773 | 2,496,699 | 13,376 |
| Yes | Unvaccinated | 5,304,173 | 1,667,534 | 1,651,342 | 823,081 | 59,447 |
|  | Primary | 16,627 | 11,141 | 2,925 | 5,123 | 0 |
|  | Primary-waned | 1,370,411 | 4,162,780 | 1,187,676 | 499,112 | 0 |
|  | Boosted | 17,001 | 97,489 | 661,915 | 542,289 | 120,380 |
|  | Booster-waned | 349,676 | 9,345,807 | 4,775,318 | 2,810,973 | 240,759 |
|  | Bivalent boosted | 6,586 | 121,171 | 250,434 | 618,184 | 120,380 |

**Metropolis-Hasting algorithm application and estimation results**

Nine parameters, $h$, $\delta^{Ⅱ}$, $\delta^{Ⅲ}$, $\delta^{Ⅳ}$, $m(t_{1})$, $m(t_{2})$, $m(t_{3})$, $m(t_{4})$, and $m(t_{5})$ are estimated using the Metropolis-Hastings algorithm, which is a Markov chain Monte Carlo method to estimate the distribution of unknown parameters by fitting to the cumulative confirmed cases of the four age groups from December 15, 2022 to May 11, 2023.^[25]^ Note that time $t_{1}$ to $t_{5}$ indicate 28-days length intervals from December 15, 2022 to May 11, 2023. We assumed Gaussian additive noise and uniform prior distribution and sampled parameters as follows:

At iteration $i$ with $x_{i}$,

1. Generate $x^{*}$ from a proposal distribution, $x^{*}\sim q(x_{i})$, $q$ is Gaussian

2. Accept or reject with acceptance rate $\alpha=min \left[ 1,\frac{f\left( x^{*}|Y \right)}{f\left( x_{i}|Y \right)} \right]$

Here, the likelihood function is

$$f\left( x_{i}|Y \right)=\int P\left( Y | x,\sigma_{i} \right)P\left( \sigma_{i} \right)d\sigma_{i}\propto\left\| Y-F\left( x_{i} \right) \right\|^{- \frac{m}{2}},$$

where$F\left( x \right)$ is the model output, $\int\sum_{i} \sum_{x} \rho^{i}\alpha I_{x}^{i} dt$, $Y$ is the data with $m$ points, and $P$ is the prior. Detailed description is in the supporting information of the study of Clarke.^[26]^ **Supplementary Figure 1.** displays sampling results. Median values [95% C.I.] of $h$, $\delta^{Ⅱ}$, $\delta^{Ⅲ}$, $\delta^{Ⅳ}$, $m(t_{1})$, $m(t_{2})$, $m(t_{3})$, $m(t_{4})$, and $m(t_{5})$ are 0·35 [0·34, 0·36], 0·97 [0·95, 1·00], 2·10 [2·05, 2·15], 6·88 [6·80, 6·97], 0·49 [0·46, 0·53], 0·19 [0·14, 0·27], 0·82 [0·66, 0·94], 0·50 [0·34, 0·65], and 0·57 [0·40, 0·79], respectively. Uncertainty of the parameter estimation is provided from the distributions of samples.

**Supplementary Figure 1.** Metropolis-Hasting algorithm application results.

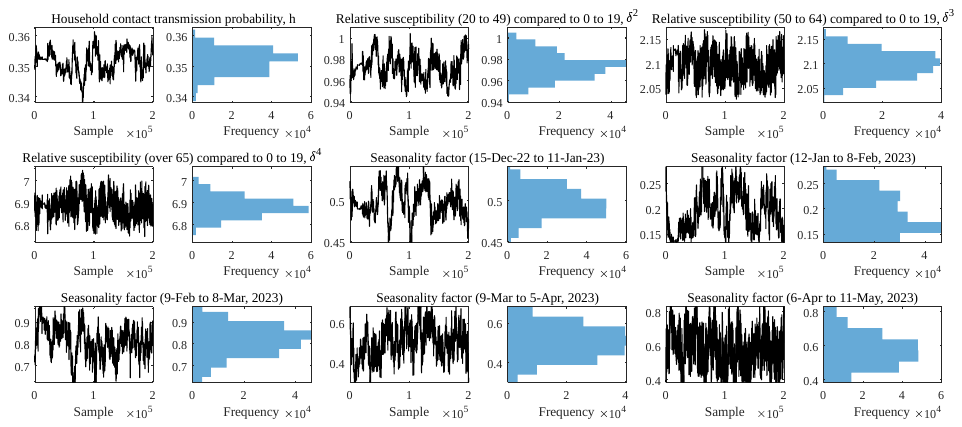

**Scenario-based study results**

Miscellaneous results which were not shown in the main manuscript are shown in this section. **Supplementary Figure 2.** shows the comparison among simulation results using different vaccination target age, with the best timing to minimize the peak size of administered severe patients. **Supplementary Figure 3.** shows the identical outline of **Figure 3.** in the main manuscript but considering different scenario, extra doses for other adults while fixing 5 million to age over 65. **Supplementary Figure 4.** shows the case that the vaccination was initiated right after the start of the extended simulation. In this case, both outbreak outcome could be similar to the results from the simulation with lesser amount of vaccine doses. **Supplementary Figure 5.** displays the time-dependent simulation results depending on the initial timing of vaccination and outbreak trend. **Supplementary Figure 6.** has same structure of **Figure 4.** in the main manuscript, while it shows the results fixing the vaccination target as age over 65 but varying the number of doses.

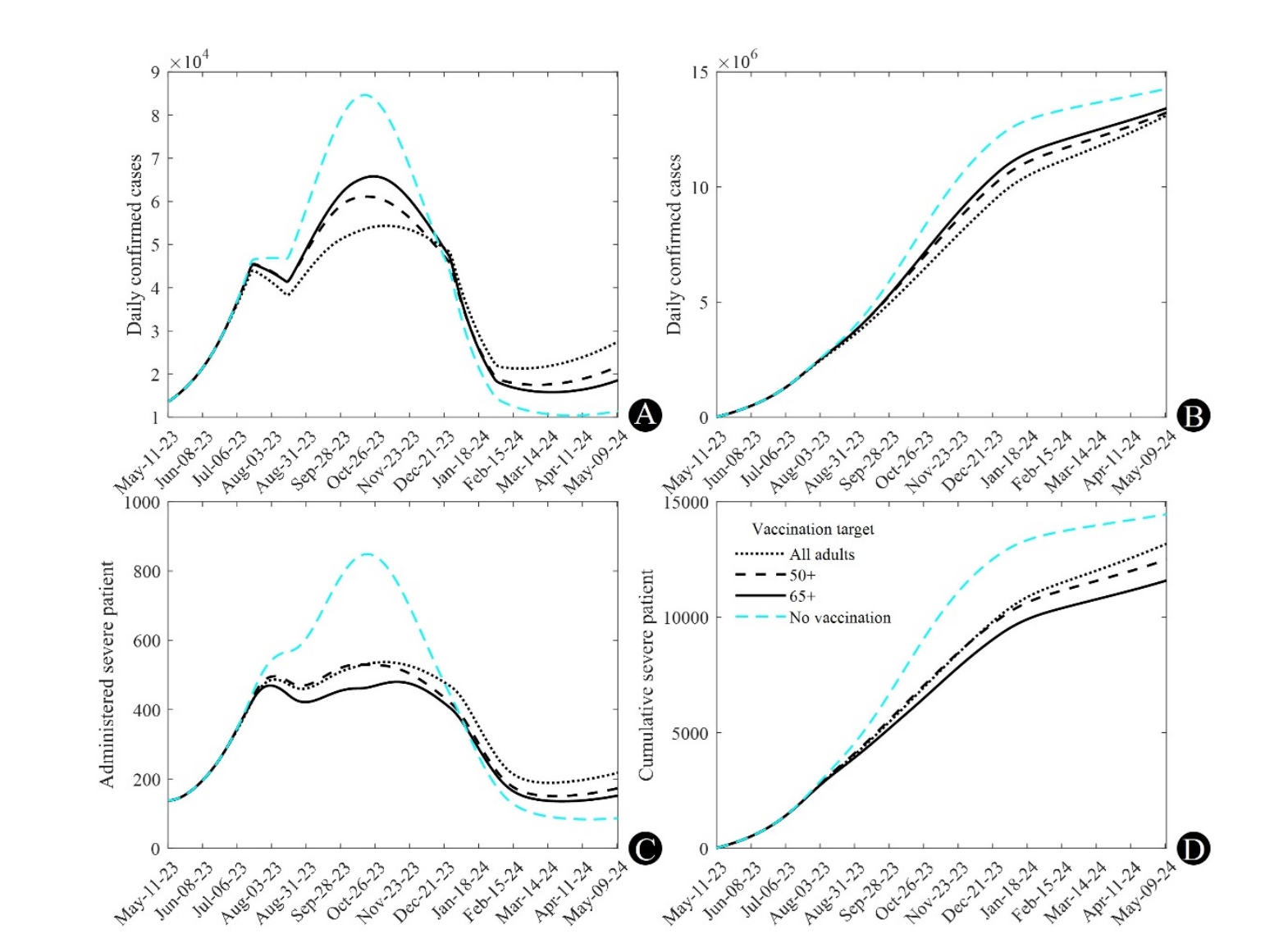

**Supplementary Figure 2.** Comparison of the scenario-based simulation results. Dotted, dashed, and solid curves indicate the scenario with vaccination target setting as all adults, age over 50, and age over 65, respectively. (A) Daily confirmed cases. (B) Cumulative confirmed cases. (C) Administered severe patients. (D) Cumulative severe cases.

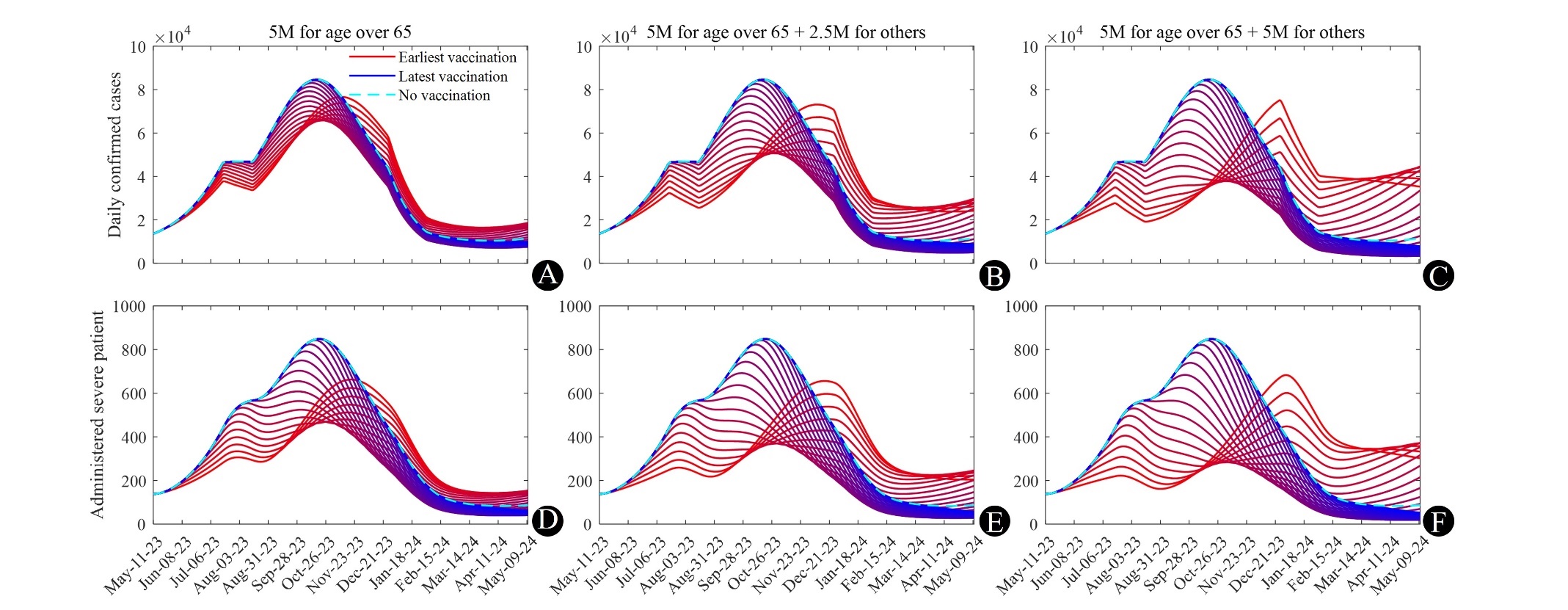

**Supplementary Figure 3.** Time series extension simulation results, (A) to (C) are daily confirmed cases and (D) to (F) are administered severe patients, fixing 5 million doses for age over 65 and setting different extra doses for other adults and timing of the vaccination. Dashed cyan curves indicate simulation without vaccination, and other curves are results of simulation with vaccination. The closer the color of the graph is to red, the earlier the vaccination time is, and the closer it is to blue, the later the vaccination is.

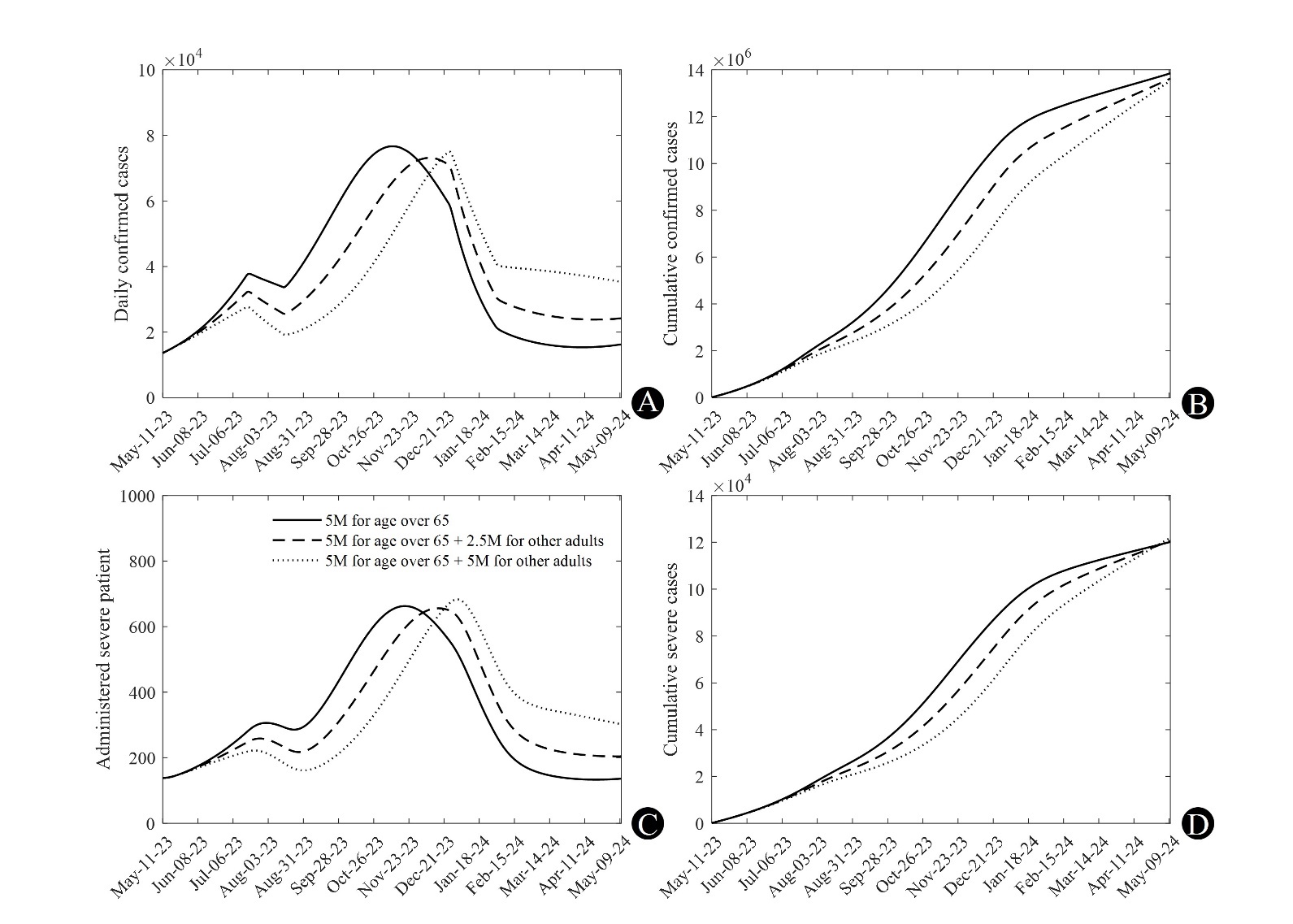

**Supplementary Figure 4.** Sampled simulation results of **Supplementary Figure 3.**, when the vaccination initiates from the beginning of simulation. Age over 65 receive 5 million doses, and solid, dashed, and dotted lines indicate extra vaccination doses for other adults as 0, 2·5 million, and 5 million, respectively. (A) Cumulative confirmed cases, (B) Cumulative severe cases, (C) Daily confirmed cases, (D) Administered severe patients.

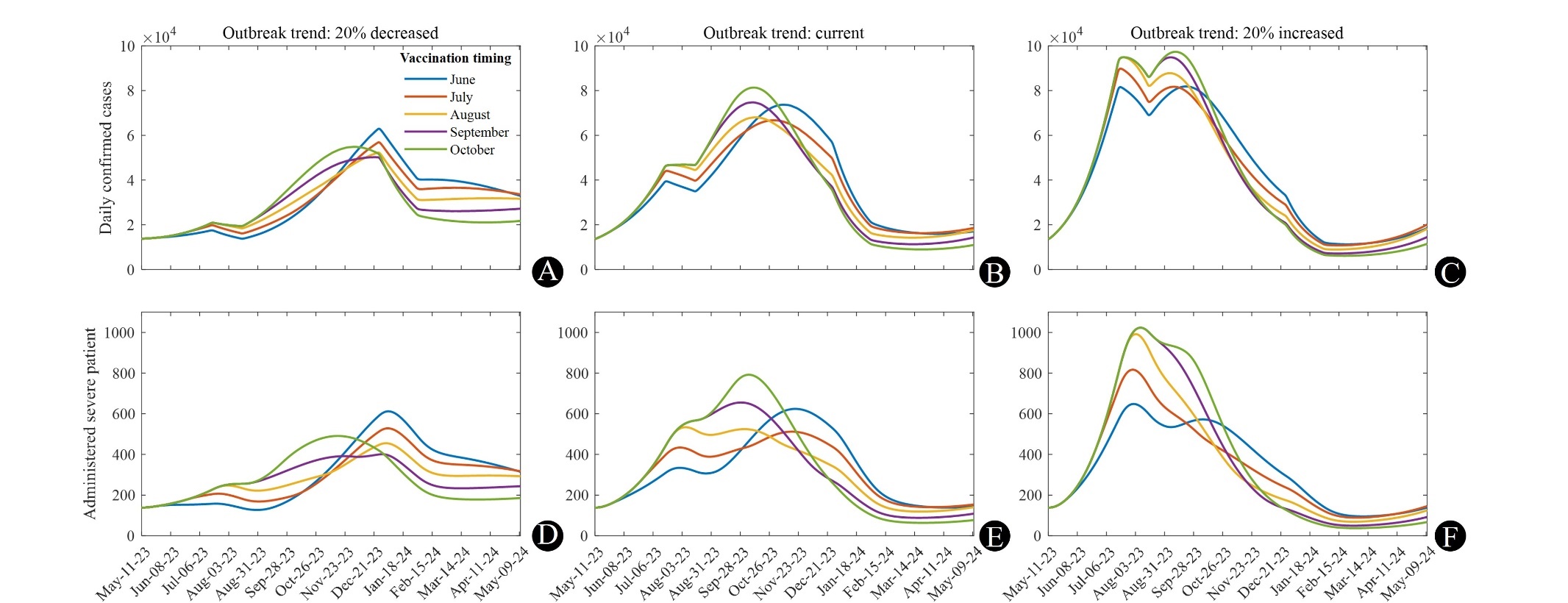

**Supplementary Figure 5.** Time series extension simulation results, (A) to (C) are daily confirmed cases and (D) to (F) are administered severe patients, fixing 5 million doses for age over 65 and setting different outbreak trend and timing of the vaccination. Colors of curve indicate the initial timing of vaccination. Note that vaccination begins from the first day of each month.

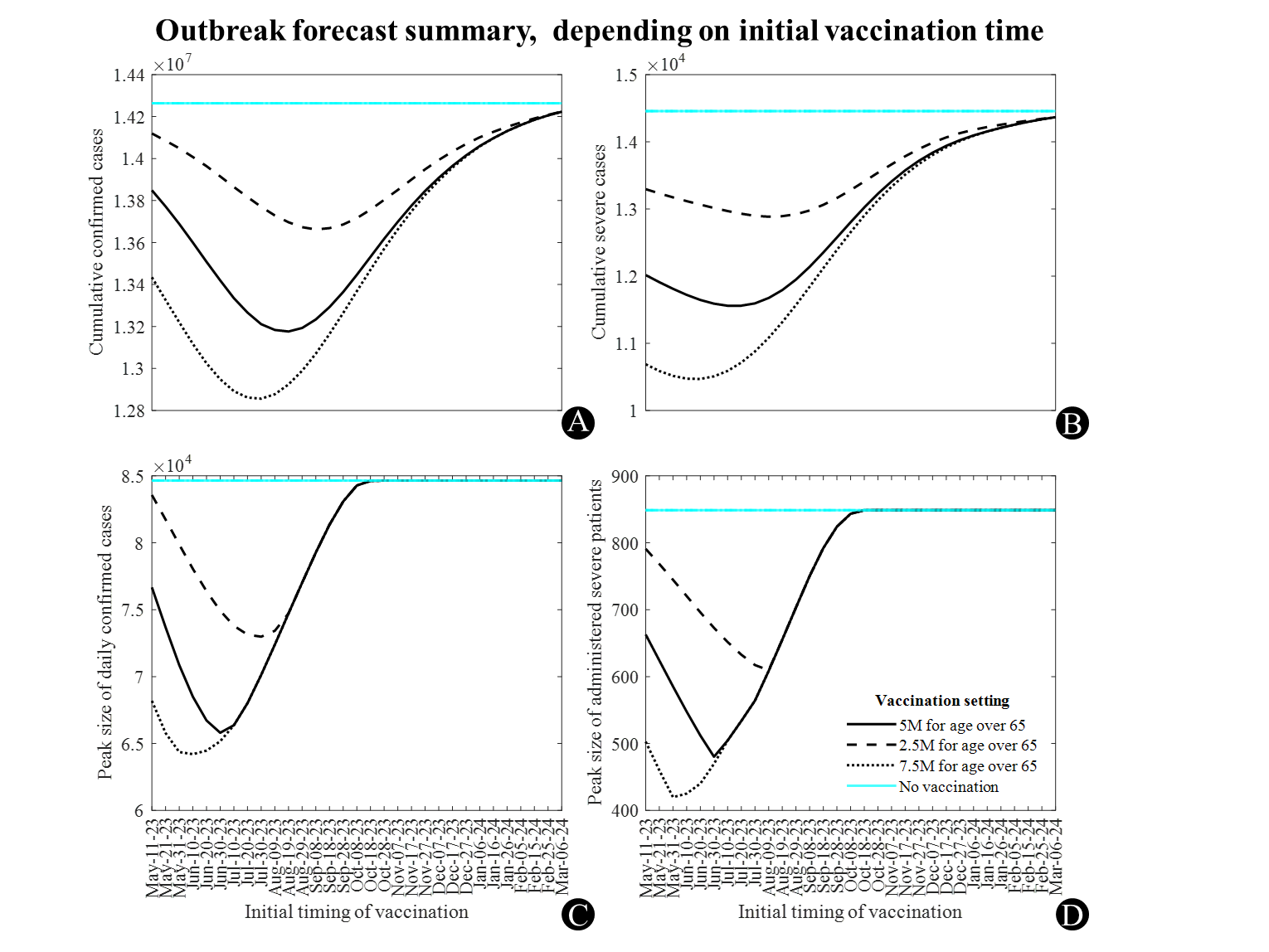

**Supplementary Figure 6.** Extended simulation results with different vaccination doses and timing, fixing the target as age over 65. (A) and (B) ((C) and (D)) shows cumulative confirmed cases and severe cases (Peak size of daily confirmed cases and administered severe patients), respectively. Note that x-axis indicates initial timing of vaccination.

**References**

1. Daily vaccine administration status [Internet]. Korea Disease Control and Prevention Agency; Available from: https://ncv.kdca.go.kr/vaccineStatus.es?mid=a11710000000 (in Korean), retrieved on May 11, 2023
2. Vaccine administration status by age [Internet]. Korea Disease Control and Prevention Agency; Available from: https://ncv.kdca.go.kr/allAgerateStatus.es?mid=a11718000000 (in Korean), retrieved on May 11, 2023
3. Ko Y, Mendoza VM, Seo Y, Lee J, Kim Y, Kwon D, Jung E. Quantifying the Effects of Non-Pharmaceutical and Pharmaceutical Interventions Against Covid-19 Epidemic in the Republic of Korea: Mathematical Model-Based Approach Considering Age Groups and the Delta Variant. Mathematical Modelling of Natural Phenomena. 2022;17:39.
4. Zee ST, Kwok LF, Kee KM, Fung LH, Luk WP, Chan TL, et al. Impact of COVID-19 Vaccination on Healthcare Worker Infection Rate and Outcome during SARS-CoV-2 Omicron Variant Outbreak in Hong Kong. Vaccines. 2022;10(8):1322.
5. Cheng HY, Jian SW, Liu DP, Ng TC, Huang WT, Lin HH. Contact tracing assessment of COVID-19 transmission dynamics in Taiwan and risk at different exposure periods before and after symptom onset. JAMA internal medicine. 2020;180(9):1156-63.
6. Payne DC, Smith-Jeffcoat SE, Nowak G, Chukwuma U, Geibe JR, Hawkins RJ, et al. SARS-CoV-2 infections and serologic responses from a sample of US Navy service members—USS Theodore Roosevelt, April 2020. Morbidity and Mortality Weekly Report. 2020;69(23):714.
7. Liu Y, Zhao S, Ryu S, Ran J, Fan J, He D. Estimating the incubation period of SARS-CoV-2 Omicron BA. 1 variant in comparison with that during the Delta variant dominance in South Korea. One Health. 2022 Dec 1;15:100425
8. Jansen L, Tegomoh B, Lange K, Showalter K, Figliomeni J, Abdalhamid B, Iwen PC, Fauver J, Buss B, Donahue M. Investigation of a SARS-CoV-2 B. 1.1. 529 (omicron) variant cluster—Nebraska, November–December 2021. Morbidity and Mortality Weekly Report. 2021;70(51-52):1782.
9. Del Águila-Mejía J, Wallmann R, Calvo-Montes J, Rodríguez-Lozano J, Valle-Madrazo T, Aginagalde-Llorente A. Secondary Attack Rate, Transmission and Incubation Periods, and Serial Interval of SARS-CoV-2 Omicron Variant, Spain. Emerging Infectious Diseases. 2022;28(6):1224.
10. Xin H, Li Y, Wu P, Li Z, Lau EH, Qin Y, Wang L, Cowling BJ, Tsang TK, Li Z. Estimating the latent period of coronavirus disease 2019 (COVID-19). Clinical Infectious Diseases. 2022;74(9):1678-81.
11. Shim E, Choi W, Song Y. Clinical Time Delay Distributions of COVID-19 in 2020–2022 in the Republic of Korea: Inferences from a Nationwide Database Analysis. Journal of Clinical Medicine. 2022;11(12):3269.
12. Hakki S, Zhou J, Jonnerby J, Singanayagam A, Barnett JL, Madon KJ, Koycheva A, Kelly C, Houston H, Nevin S, Fenn J. Onset and window of SARS-CoV-2 infectiousness and temporal correlation with symptom onset: a prospective, longitudinal, community cohort study. The Lancet Respiratory Medicine. 2022;10(11):1061-73.
13. COVID-19 domestic occurrence status [Internet]. Ministry of Health and Welfare of South Korea; Available from: https://ncov.kdca.go.kr/bdBoardList_Real.do?brdId=1&amp;brdGubun=11&amp;ncvContSeq=&amp;contSeq=&amp;board_id=&amp;gubun= (in Korean), retrieved on May 11, 2023
14. Vaccine effectiveness against severity and fatality [Online]. Korea Disease Control and Prevention Agency; Available from: https://www.kdca.go.kr/board/board.es?mid=a20602010000&bid=0034&list_no=716913&act=view
15. COVID-19 response weekly news, 2022 Mar 11 [Internet]. Seoul Metropolitan Government; Available from: https://www.seoul.go.kr/seoulcom/fileDownload.do?fileName=corona/daily-news-review_220311_507.pdf (in Korean), retrieved on May 11, 2023
16. Patient management: quarantine period, release criteria, and evidence [Internet]. Korea Disease Control and Prevention Agency; Available from: https://ncv.kdca.go.kr/hcp/page.do?mid=0302 (in Korean), retrieved on May 11, 2023
17. Jo S, Nam HK, Kang H, Cho SI. Associations of symptom combinations with in-hospital mortality of coronavirus disease-2019 patients using South Korean National data. Plos one. 2022;17(8):e0273654.
18. Altarawneh HN, Chemaitelly H, Ayoub HH, Tang P, Hasan MR, Yassine HM, Al-Khatib HA, Smatti MK, Coyle P, Al-Kanaani Z, Al-Kuwari E. Effects of previous infection and vaccination on symptomatic omicron infections. New England Journal of Medicine. 2022;387(1):21-34.
19. Tseng HF, Ackerson BK, Luo Y, Sy LS, Talarico CA, Tian Y, Bruxvoort KJ, Tubert JE, Florea A, Ku JH, Lee GS. Effectiveness of mRNA-1273 against SARS-CoV-2 Omicron and Delta variants. Nature Medicine. 2022;28(5):1063-71.
20. Andrews N, Stowe J, Kirsebom F, Toffa S, Rickeard T, Gallagher E, Gower C, Kall M, Groves N, O’Connell AM, Simons D. Covid-19 vaccine effectiveness against the Omicron (B. 1.1. 529) variant. New England Journal of Medicine. 2022;386(16):1532-46.
21. Kirsebom FC, Andrews N, Stowe J, Toffa S, Sachdeva R, Gallagher E, Groves N, O'Connell AM, Chand M, Ramsay M, Bernal JL. COVID-19 vaccine effectiveness against the omicron (BA. 2) variant in England. The Lancet Infectious Diseases. 2022;22(7):931-3.
22. Bobrovitz N, Ware H, Ma X, Li Z, Hosseini R, Cao C, Selemon A, Whelan M, Premji Z, Issa H, Cheng B. Protective effectiveness of previous SARS-CoV-2 infection and hybrid immunity against the omicron variant and severe disease: a systematic review and meta-regression. The Lancet Infectious Diseases. 2023;23(5):556-7.
23. Population status by age [Internet]. Ministry of the Interior and Safety; Available from: https://jumin.mois.go.kr/ageStatMonth.do (in Korean), retrieved on May 11, 2023
24. Healthcare worker status [Internet]. Statistics Korea; Available from: https://kosis.kr/statHtml/statHtml.do?orgId=101&tblId=DT_2KAAC01 (in Korean), retrieved on May 11, 2023
25. Hastings WK. Monte‐Carlo sampling methods using Markov chains and their applications. Biometrika. 1970;57:97‐109.
26. Andrejko KL, Pry JM, Myers JF, Fukui N, DeGuzman JL, Openshaw J, Watt JP, Lewnard JA, Jain S, COVID C, COVID C. Effectiveness of face mask or respirator use in indoor public settings for prevention of SARS-CoV-2 infection—California, February–December 2021. Morbidity and Mortality Weekly Report. 2022;71(6):212.
